# Gaps in Congenital Heart Disease Care: Social Drivers and Clinical Consequences

**DOI:** 10.64898/2026.06.24.26356504

**Authors:** Abbas H. Zaidi, Adam Alberts, Abraham Kwan, Gumpili Sai Prashanthi, Kathy Jenkins, Susan F. Saleeb, Erica Sood, Anne Kazak, Sarah Deferranti

**Affiliations:** Nemours Cardiac Center, Nemours Children’s Health, Wilmington, DE; Department of Pediatrics, Sidney Kimmel Medical College, Thomas Jefferson University, Philadelphia, PA; Department of Pediatrics, Nemours Children’s Health, Wilmington, DE; Department of Internal Medicine, ChristianaCare Hospital, Wilmington, DE; Division of Ambulatory Cardiology, Department of Cardiology, Boston Children’s Hospital, Boston, MA; Department of Pediatrics, Harvard Medical School, Boston, MA; Nemours Children’s Health Center for Healthcare Delivery Science, Wilmington, DE

**Author notes:** Correspondence: Abbas H. Zaidi, MD; Nemours Cardiac Center, Nemours Children’s Health; 1600 Rockland Road, Wilmington, Delaware, 19803.

**Keywords:** congenital heart disease, gaps in care, loss to follow-up

## Abstract

**Background:** Gaps in care (GIC) among patients with congenital heart disease (CHD) are associated with adverse outcomes, yet the specific social and healthcare-related factors contributing to GIC and the clinical consequences of delayed re-engagement in care remain poorly characterized. Large electronic medical record datasets often cannot distinguish true GIC from clinically appropriate care patterns or capture the patient-level factors contributing to GIC.

**Methods:** We conducted a retrospective cohort study, combining large data with manual chart review, of 1,746 patients of all ages with surgically repaired CHD between 2003 and 2020 at a tertiary care center serving four states. GIC was defined as more than 3 years and 3 months between cardiology visits and exceeding the physician recommended follow-up interval.

**Results:** Of the cohort, 916 patients (52%) met criteria for potential GIC. Following a structured manual chart review, a substantial subset was reclassified as having appropriate care, leaving 275 patients (15.7%) with true GIC. After multivariable adjustment, older age and simple anatomic CHD complexity were independently associated with GIC. Among patients with GIC, 17.8% had a documented contributor, most commonly insurance instability or social factors. Forty-one and one-half percent returned to care (RTC), and many were asymptomatic but had significant disease progression. Thirteen percent of patients who RTC required cardiac intervention, including semi-urgent or urgent procedures, and 26.7% of those requiring intervention experienced significant morbidity or mortality, including stroke, infective endocarditis, urgent transplant referral, or death. These outcomes occurred across all levels of CHD complexity, including patients with simple CHD.

**Conclusions:** GIC remain prevalent in patients with surgically repaired CHD and are associated with significant morbidity and mortality across the full spectrum of anatomic complexity. They are most often driven by insurance instability and social vulnerability rather than clinical factors, and many adverse outcomes may be preventable with consistent longitudinal care. These findings support a shift toward proactive care models that integrate standardized follow-up pathways, systematic assessment of patient-level needs, and emerging analytic tools to identify at-risk patients before GIC occur.

**CLINICAL PERSPECTIVE:** *What Is New?:*

- Gaps in care (GIC) remained common among patients with surgically repaired CHD and were associated with insurance instability, social vulnerability, asymptomatic disease progression, major adverse clinical events, and death.

*What Are the Clinical Implications?:*

- Reducing GIC and its consequences will require systematic assessment of social and patient-reported needs, education that asymptomatic status does not imply disease stability, and emerging predictive tools such as artificial intelligence to identify patients at risk for disengagement before adverse outcomes occur.

## Introduction

Advances in surgical and medical management have significantly improved survival among patients with congenital heart disease (CHD), resulting in a growing population requiring lifelong longitudinal care.^1–7^ Ongoing follow-up is essential to monitor for residual lesions, late complications, and CHD progression.^4–7^ Despite this need, gaps in care (GIC) remain common and represent a persistent challenge in CHD management.^8–16^

Over the last decade, a growing body of research has emphasized the impact of social drivers of health (SDOH) across the continuum of CHD, shaping early diagnosis, patterns of disease burden, infant mortality, postoperative recovery, and ongoing engagement with care, including adherence to follow-up care.^9,17–24^ Studies have shown that GIC are associated with age, disease complexity, and sociodemographic factors such as race, ethnicity, and insurance status, with SDOH consistently contributing to barriers in care continuity.^8,11,13,25–27^ Despite growing recognition of GIC for CHD, important limitations remain in the existing literature.^8,13,25^ Most studies relied on administrative definitions or structured electronic medical record (EMR) without manual review to identify GIC, which may misclassify patients by failing to distinguish true discontinuity from clinically appropriate care, such as follow-up care at outside institutions or provider-recommended intervals longer than the GIC definition used in studies. In addition, while SDOH and health system factors associated with GIC have been described, there remains a limited understanding of the patient-level drivers, including the specific insurance and social circumstances that disrupt care continuity in real-world settings.^8,11,13,25–27^

Equally important, the downstream clinical consequences of GIC are not well characterized. Existing studies demonstrated associations between loss to follow-up (LTFU) care and adverse outcomes, but lack granular data describing what occurred during periods of disengagement or at the time of return to care (RTC).^12,28–32^ In particular, there is minimal insight into clinical trajectories at re-engagement, including the timing and urgency of required interventions and the extent to which GIC contribute to morbidity and mortality across the spectrum of CHD complexity.^4,10–12,32^

To address these gaps, this study integrates structured EMR data with detailed manual chart review to (1) distinguish ongoing GIC, (2) identify and characterize patient-level contributors to GIC, and (3) describe clinical trajectories following GIC, including patterns of RTC, need for interventions, and associated morbidity and mortality.

## Methods

### Study Design and Patient Selection

We conducted a retrospective cohort study of patients with CHD who underwent cardiac surgery at a pediatric subspecialty hospital serving Delaware, Pennsylvania, New Jersey, and Maryland between January 2003 and May 2020. Institutional review board approval was obtained before data collection, and informed consent was waived due to the retrospective nature of the study. This study was approved by the Nemours Children’s Health Institutional Review Board with a waiver of informed consent due to the retrospective nature of the study.

The study period was selected to align with the institution’s transition to a unified EMR system and to ensure all patients had at least 3 years of potential follow-up care at the time of analysis to allow accurate identification of GIC.

Inclusion criteria consisted of patients with surgically repaired CHD within the institution’s tertiary cardiothoracic surgical program who required at least one outpatient cardiology visit beyond the standard postoperative surgical follow up, to ensure continuity of care at the institution. The Society of Thoracic Surgeons database was used to identify the initial cohort of patients with surgically intervened CHD. Each patient’s cardiac diagnosis was mapped to an anatomic severity classification of simple, moderate, or complex based on established American College of Cardiology/American Heart Association (ACC/AHA) guidelines.^4,5,8,13^

Exclusion criteria included patients who did not undergo cardiac surgery at the tertiary care institution or those without CHD but needed a cardio-thoracic surgery, such as those requiring extracorporeal membrane oxygenation for non-congenital indications, and those undergoing non-cardiac thoracic surgery (e.g., pulmonary embolectomy).

A clinically significant “ongoing GIC” was defined as a lapse of more than 3 years and 3 months between cardiology visits or exceeded the physician recommended follow-up interval documented at the prior cardiology visit by more than 3 months to allow room for clinical practice variability based on CHD complexity and potential scheduling related issues.

### Data Sources and Variable Extraction Sociodemographic and Clinical variables

Sociodemographic variables included race and ethnicity, categorized into White versus non-White and Hispanic versus non-Hispanic, respectively. Preferred language was categorized as English or non-English. State of residence was recorded. Insurance-related variables included insurance status categorized as public (Medicaid, Medicare, managed Medicaid, Tricare) or private, insurance composition over time (public only, private only, or mixed), and number of unique payors across the patient’s history (0, 1, or ≥ 2). Additional utilization-related variables included whether the patient had ever no-showed to a visit, number of listed guardians, total number of encounters, total number of in-person encounters, total number of telephone encounters where the calls were made either by patient or the healthcare team (e.g., scheduling, nursing etc), and average copay across visits.

### Geographic and Neighborhood-Level Measures

The distance from the patient’s residence, based on zip code, to the tertiary care hospital or its nearest affiliated cardiology site was calculated. These zip codes also were used to assign Child Opportunity Index (COI) scores, using COI 3.0, which incorporates 44 indicators across education, health, environment, and social and economic domains.^8,13,33^ Scores were categorized into standard groupings, and domain-specific analyses were performed.

### Manual Chart Review and Classification of Ongoing GIC

A structured manual chart review was performed for all patients with potential “ongoing GIC” to determine whether the observed interval represented ongoing GIC or an appropriate and justifiable care pattern (e.g., moved care to another institution) not captured in structured data. Manual chart review was performed by author AA and reviewed and discussed bi-weekly via meetings with AZ. This chart review included detailed encounter histories across outpatient visits, inpatient encounters, telephone encounters, imaging studies, and outside hospital medical records when available.

The chart review followed a sequential and standardized process. First, the cardiology clinic visit immediately preceding the assumed GIC was reviewed to determine the physician-recommended follow-up interval. Second, the full clinic note was reviewed to identify documentation of potential contributing factors to the GIC, including SDOH, symptoms, and care recommendations. Third, additional cardiology documentation was reviewed, including telephone encounters, canceled appointments, cardiac conference notes, imaging reports, and internal communications, including scheduling documentation and non-permanent notes used to track outreach efforts. Fourth, external data were reviewed using available cross-institution data sharing tools (“Care Everywhere” tool in EMR, scanned notes, etc.) to determine whether the patient was receiving cardiology care at another institution. Fifth, non-cardiac visits within the health system, including primary care and subspecialty encounters, were reviewed to identify evidence of care continuity elsewhere or contextual explanations for the absence of cardiology follow-up care. Sixth, social work encounters were reviewed when available.

Patients were classified as having “no GIC” if they had a justifiable cause e.g., if they were followed at an outside institution, formally discharged from cardiology care, had provider-recommended follow-up care exceeding 3 years, or were deceased with appropriate follow-up prior to death. Patients without any identifiable justification after this review were classified as having “true GIC”.

### Classification of Contributors to GIC

Among patients classified as having true GIC, chart review was used to determine whether contributing factors were documented. These contributors were further sub-classified after reading contextual details through the EMR notes outlined above. If multiple contributing factors were present, the predominant contributor was assigned based on documentation. If no contributing factor was identified despite comprehensive review, the patient was classified as having no documented reason for ongoing GIC.

### Care Trajectories and Reasons for RTC

After chart review, patients with true GIC were further classified as LTFU (did not return for a visit since GIC) or as patients who RTC (returned to clinical care after GIC). For patients who RTC, the index return visit was reviewed to determine the reason for re-engagement. For patients who RTC, reasons for the GIC, cause/purpose of re-engagement, detailed clinical trajectories including need for re-intervention (surgical versus cardiac catheterization), and any significant morbidity or mortality were recorded.

### Classification of Interventions, Urgency, and Outcomes

For patients who RTC and required cardiac interventions, all subsequent cardiology visits including inpatient and outpatient, imaging studies, diagnostic tests, intervention and surgical procedure notes were reviewed to determine management course and outcomes. Interventions were classified based on urgency. Elective interventions were defined as those that could be completed beyond 6 months. Semi-urgent interventions were defined as those requiring completion within 6 months due to disease progression or concern for worsening clinical status. Urgent interventions were defined as those occurring in the setting of emergency department presentation or hospitalization due to hemodynamic compromise or acute clinical deterioration. Clinical outcomes, including morbidity and mortality, were identified through review of inpatient records, procedural documentation, and follow-up encounters.

## Statistical Analysis

Baseline demographic, clinical, insurance, and SDOH variables were summarized for the overall cohort and then stratified by the presence or absence of an ongoing GIC. Continuous variables were assessed using the Shapiro–Wilk test and reported as medians with interquartile ranges. Categorical variables were presented as frequencies. Non-normally distributed continuous variables were compared using the Mann–Whitney U test. Categorical variables were compared using Chi-Square χ² tests. Variables evaluated included age, sex, anatomic CHD complexity, race, ethnicity, primary language, state of residence, insurance status and composition, number of insurance changes, number of listed guardians, distance to care, and COI overall and domain-specific categories.

A multivariable logistic regression model was constructed to identify factors associated with ongoing GIC. Variables included in the model were selected based on the literature and included age, sex, anatomic CHD severity, race, ethnicity, primary language, state of residence, insurance characteristics, distance to care, and COI measures.^8,13^ Results are reported as odds ratios with corresponding 95% confidence intervals and p-values. Statistical significance was defined as a two-sided p-value < 0.05. All statistical analyses were performed using Python (Python Software Foundation) and R version 4.2.3.

## Results

### Cohort Characteristics

A total of 1,746 patients with surgically repaired CHD between 2003 and 2020 residing in Delaware, Pennsylvania, New Jersey, or Maryland were included in the cohort (Table 1). The median age was 9.53 years (range 3.98-15.33), 47.3% were female, and 53.4% were White non-Hispanic. Most patients resided in Delaware (38.3%) or Pennsylvania (38.3%), with the remaining patients residing in New Jersey or Maryland at their visit prior to GIC.

**Table 1.** Baseline characteristics of patients with CHD stratified by ongoing GIC.

| **Variable N (%)** | **Total** | **GIC** | **No GIC** | **p value** |
| --- | --- | --- | --- | --- |
| **Total** | 1746 (100) | 275 (15.7) | 1471 (84.3) | < 0.001 |
| **Age** | 9.53 (3.98-15.33) | 10.97 (6.51-16.48) | 8.71 (3.6-14.79) | < 0.001 |
| Q1 (0.0033, 8.712] | 833 (47.7) | 96 (34.9) | 737 (50.1) |  |
| Q2 (8.712, 17.386] | 629 (36.0) | 125 (45.5) | 504 (34.3) |  |
| Q3 (17.386, 26.061] | 253 (14.5) | 50 (18.2) | 203 (13.8) |  |
| Q4 (26.061, 34.735] | 31 (1.8) | 4 (1.5) | 27 (1.8) |  |
| **Female Gender** | 826 (47.3) | 134 (48.7) | 692 (47.0) | 0.6372 |
| **Cardiac Anatomy** |  |  |  |  |
| Simple | 560 (32.1) | 135 (49) | 425 (29.0) | < 0.001 |
| Moderate | 717 (41.1) | 117 (42.5) | 600 (40.7) | < 0.001 |
| Complex | 469 (26.8) | 23 (8.5) | 446 (30.3) | < 0.001 |
| **English speaking (n=1633)** | 1508 (92.3) | 223 (89.9) | 1285 (92.8) | 0.1525 |
| **State of residence** |  |  |  | 0.0005 |
| DE | 668 (38.3) | 110 (40.0) | 558 (38.0) |  |
| MD | 69 (3.9) | 15 (5.4) | 54 (3.7) |  |
| NJ | 341 (19.5) | 72 (26.2) | 269 (18.2) |  |
| PA | 668 (38.3) | 78 (28.4) | 590 (40.1) |  |
| **Distance to main hospital (miles)**<br>**(n=1744)** | 28.64 (16.51-51.69) | 29.52 (16.51-53.21) | 27.84 (16.51-48.44) | 0.2261 |

| **Variable N (%)** | **Total** | **GIC** | **No GIC** | **p value** |
| --- | --- | --- | --- | --- |
| **Distance to closest clinic (miles)** | 9.35 (5.34-21.19) | 9.31 (5.36-15.22) | 9.41 (5.34-22.74) | 0.1597 |
| **<2 Guardians** | 444 (25.4) | 81 (29.9) | 363 (24.7) | 0.1108 |
| **<u>Insurance History</u>** |  |  |  |  |
| **On public Insurance** | 1035 (59.3) | 186 (67.6) | 849 (57.7) | 0.0026 |
| Private only | 652 (37.3) | 88 (32) | 564 (38.3) | 0.0021 |
| Public only | 728 (41.7) | 108 (39.3) | 620 (42.2) | 0.0021 |
| Mixed Insurance (public and private) | 366 (21.0) | 79 (28.7) | 287 (19.5) | 0.0021 |
| Number of Insurances |  |  |  | 0.0011 |
| 0 | 383 (21.9) | 37 (13.4) | 346 (23.5) |  |
| 1 | 872 (49.9) | 152 (55.3) | 720 (49.0) |  |
| 2+ | 491 (28.2) | 86 (31.3) | 405 (27.5) |  |
| **Race and Ethnicity (n=1706)** |  |  |  | 0.0071 |
| Non-White Hispanic | 105 (6.2) | 28 (10.3) | 77 (5.3) |  |
| Non-White non-Hispanic | 507 (29.7) | 95 (35.2) | 412 (28.7) |  |
| White Hispanic | 40 (2.3) | 6 (2.2) | 34 (2.4) |  |
| White non-Hispanic | 911 (53.4) | 123 (45.) | 788 (54.9) |  |
| **COI Overall** |  |  |  | 0.0938 |
| High/very high | 824 (47.2) | 117 (42.5) | 707 (48.1) |  |
| Moderate | 453 (25.9) | 70 (25.5) | 383 (26) |  |

| Variable N (%) | Total | GIC | No GIC | p value |
| --- | --- | --- | --- | --- |
| Very low/low | 469 (26.9) | 88 (32) | 381 (25.9) |  |
| **COI Education** |  |  |  | 0.2379 |
| High/very high | 805 (46.1) | 122 (44.4) | 683 (46.43) |  |
| Moderate | 429 (24.6) | 61 (22.2) | 368 (25.0) |  |
| Very low/low | 512 (29.3) | 92 (33.4) | 420 (28.55) |  |
| **COI Health & Environment** |  |  |  | 0.9224 |
| High/very high | 766 (43.9) | 120 (43.7) | 646 (43.9) |  |
| Moderate | 425 (24.3) | 65 (23.6) | 360 (24.5) |  |
| Very low/low | 555 (31.8) | 90 (32.7) | 465 (31.6) |  |
| **COI Social & Economic** |  |  |  | 0.0162 |
| High/very high | 805 (46.1) | 106 (38.6) | 699 (47.5) |  |
| Moderate | 405 (23.2) | 68 (24.7) | 337 (22.9) |  |
| Very low/low | 536 (30.7) | 101 (36.7) | 435 (29.6) |  |

Data are presented as n (%) or median (IQR). P values were calculated using $\chi^2$ tests for categorical variables and Wilcoxon rank-sum tests for continuous variables. Percentages may not total 100% because of rounding or missing data. **Abbreviations:** CHD, congenital heart disease; GIC, gap in care; COI, Child Opportunity Index; IQR, interquartile range; DE, Delaware; MD, Maryland; NJ, New Jersey; PA, Pennsylvania.

With respect to CHD complexity, 26.8% were classified as complex, 41.1% as moderate, and 32.1% as simple. Most patients (59.3%) had public insurance, and 28.2% had multiple insurance plans over time.

### Classification of GIC and Documented Contributors

Using the predefined definition, 916 patients (52%) had intervals ≥ 3 years and 3 months between cardiology visits and were classified as having potential GIC (Figure 1). Following structured chart review, a substantial proportion of these patients were reclassified as having appropriate care, while 275 patients were identified as having true GIC as shown in Figure 1.

**Figure 1.**
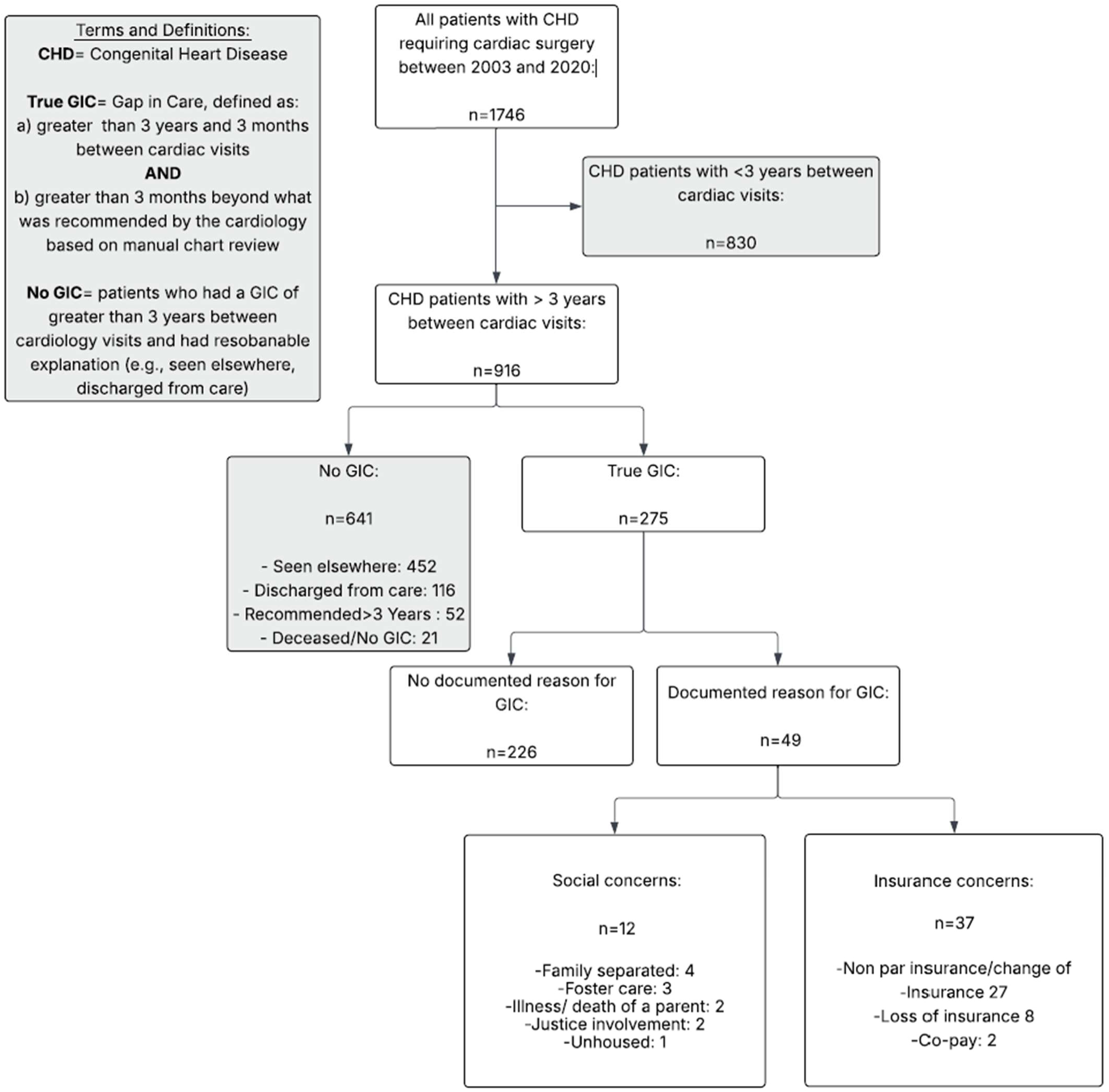
Cohort derivation and classification of GIC among patients with CHD.

Among patients with ongoing GIC, 49 (17.8%) had a documented contributing factor identified through chart review. Of these, 37 cases were attributed to insurance-related factors and 12 to social factors.

Among insurance-related contributors, the majority were due to non-participating insurance or changes in insurance coverage (72.9%, n=27), followed by loss of insurance (21.6%, n=8), and copay-related barriers (5.4%, n=2).

Among social contributors, family separation was the most frequently documented factor (33.3%, n=4), followed by foster care placement (25%, n=3), death of a family member (16.6%, n=2), and justice system involvement (16.6%, n=2).

### Care Trajectories and Reasons for RTC

Among the 114 patients who RTC, the majority (64.9%, n=74) had no documented reason for re-engagement (Figure 2). Among the 40 patients (35.1%) with a documented reason, most returned due to symptoms (72.5%, n=29). The most commonly reported presenting complaint was chest pain (37.9%, n=11), followed by exercise intolerance (20.7%, n=6), palpitations (13.8%, n=4), syncope or lightheadedness (10.3%, n=3), and non-cardiac concerns (17.2%, n=5). The remainder returned for cardiac clearance prior to non-cardiac procedures (27.5%, n=11), such as dental procedures (36.4%, n=4), non-cardiac surgery (54.5%, n=5), and sports clearance (9%, n=1) (Figure 2).

**Figure 2.**
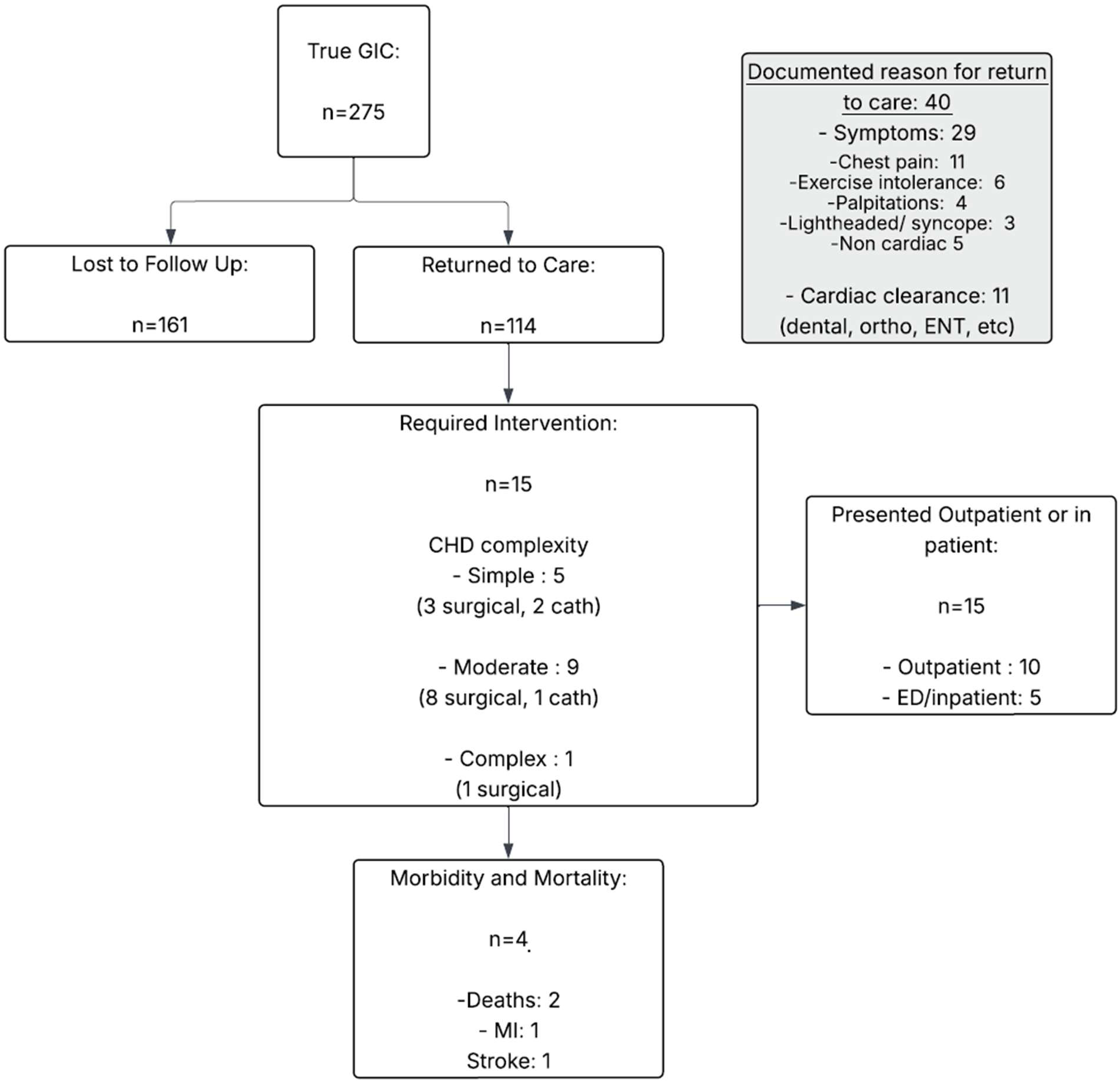
Outcomes following GIC, including RTC, interventions, and associated morbidity and mortality. **Abbreviations:** GIC, gap in care; CHD, congenital heart disease; ED, emergency department; ENT, ear, nose, and throat; MI, myocardial infarction.

### Characteristics and Clinical Trajectories of RTC Patients

Among RTC patients, 15 (13.2%) required cardiac intervention (Figure 2 and Supplemental Table S1). These interventions occurred across all levels of CHD complexity, including simple (33.3%, n=5), moderate (60%, n=9), and complex lesions (6.7%, n=1). Among those requiring intervention, four patients (26.7%) experienced significant morbidity or mortality.

Among RTC patients, there was substantial heterogeneity in CHD diagnoses, disease complexity, and clinical trajectories (Supplemental Table S1). Patients spanned the full spectrum of CHD, including simple lesions such as atrial septal defects and ventricular septal defects, as well as moderate and complex CHD, including coarctation of the aorta, atrioventricular canal defects, Shone’s complex, and single ventricle physiology. Duration of GIC prior to RTC was frequently >4 years and in some instances >10 years.

Despite this heterogeneity, several consistent patterns emerged. A subset of patients RTC in a clinically stable state and were found to have progressive structural or hemodynamic abnormalities requiring elective intervention, often in the absence of symptoms (Supplemental Table S1). In contrast, other patients presented with advanced disease, including heart failure, cyanosis, or complications such as endocarditis, requiring semi-urgent or urgent intervention. Urgent presentations frequently occurred in patients with prolonged GIC and included emergency department presentation or hospitalization.

Interventions included both catheter-based and surgical approaches and were required across all levels of CHD complexity. Importantly, significant morbidity and mortality were observed, including stroke, myocardial infarction, or death, often in the setting of delayed recognition of disease progression.

### Multivariable Analysis of Factors Associated with GIC

In the multivariable logistic regression model (Supplemental Table S2), several factors were independently associated with ongoing GIC. Older age at most recent visit was associated with increased odds of GIC (OR 1.06 per year, 95% CI 1.02–1.10, p=0.001). Compared with patients residing in Delaware, those residing in New Jersey had higher odds of GIC (OR 1.78, 95% CI 1.15–2.74, p=0.01), while residence in Maryland or Pennsylvania did not reach statistical significance. Anatomic CHD complexity demonstrated a strong inverse association with GIC, as patients with moderate (OR 0.51, 95% CI 0.36–0.71, p<0.001) or complex CHD (OR 0.13, 95% CI 0.07–0.21, p<0.001) had substantially lower odds of GIC compared to patients with simple CHD. Sex, primary language, number of guardians, distance to care, race and ethnicity, insurance characteristics, and COI domain measures were not independently associated with GIC.

## Discussion

In this study of patients with surgically repaired CHD, GIC remained common, clinically consequential, and frequently driven by social and structural factors that are difficult to anticipate through routine clinical practice. Although a stringent definition of GIC (≥3 years and 3 months between cardiology visits and exceeding the recommended follow-up interval) was used as a starting point, structured manual chart review reclassified a substantial proportion of these patients as having appropriate care pathways, leaving 275 (15.7%) with true GIC. This rate is likely a conservative estimate of the true burden of GIC, as clinically meaningful gaps also may occur among patients who remain nominally engaged yet experience delays, missed follow-up opportunities, or variability in clinician-recommended intervals that are not captured by our time-based definition. Even under this conservative definition, GIC were not benign: among patients who returned to care, 13.2% required cardiac intervention, and more than one in four of those experienced significant morbidity or mortality, including stroke from intracardiac shunting, infective endocarditis, urgent transplant referral, or death.

Classification as having an appropriate care pathway should not be interpreted as the absence of clinical risk. Patients followed at outside institutions remain vulnerable to disruptions stemming from insurance changes, referral barriers, incomplete record transfers, and loss of disease-specific context during transitions across health systems.^11,15,16^ Similarly, longer clinician-recommended follow-up intervals do not eliminate the possibility of disease progression, particularly in CHD, in which residual lesions and late complications may evolve silently. Prior literature and contemporary guideline statements, including the 2018 ACC/AHA and 2025 updated guidelines for adults with CHD, emphasize lifelong surveillance across all anatomic categories.^4,5^ Our data demonstrate that real-world practice falls short of this standard even within a tertiary care system, and that the observed adverse outcomes may reflect the consequences of particularly significant GIC.

A central finding of this study is that GIC occur across the full spectrum of CHD complexity. After multivariable adjustment, patients with moderate and complex CHD had substantially lower odds of GIC compared to patients with simple CHD, which likely is in part due to physician recommendation to follow up at a longer duration for patients with simple CHD. However in this cohort patient with simple CHD with GIC had significant morbidity related to disease progression, providing quantitative reinforcement for what has long been suspected but inconsistently emphasized: simple CHD is not inherently low risk.^13,15,25,32,34,35^ The clinical trajectories among patients with simple lesions in this cohort make this point concretely. Patients with surgically repaired ventricular septal defects, atrial septal defects, and bicuspid aortic valves returned after prolonged GIC with progressive aortic regurgitation, right heart dilation requiring surgery, mixed aortic valve disease requiring valve replacement, and, in one case, infective endocarditis with intracardiac vegetation. These outcomes challenge the perception that anatomic simplicity equates to longitudinal clinical safety and reinforces the need to maintain consistent surveillance across all surgically repaired CHD.^13,25,32^

When contributors to GIC were identifiable, they were dominated by insurance instability and social vulnerability. These factors point to a particularly fragile subgroup in whom social and structural conditions are the proximate determinants of both care continuity and clinical outcomes.^8,11,13,26,27^ Importantly, these contributors were dynamic, often arose between rather than during clinical encounters, and were inconsistently documented, suggesting the true burden of insurance and social barriers is substantially greater than what was captured. The state-level finding from the adjusted analysis – patients residing in New Jersey had higher odds of GIC than those in Delaware – further suggests that geographic variation in insurance networks, referral pathways, and access to in-network subspecialty care contribute to GIC in ways that traditional SDOH frameworks may not fully capture. Taken together, these findings argue that addressing GIC requires health systems to move beyond passive documentation of social risk and toward processes that actively identify, monitor, and respond to evolving patient circumstances in real time.

Among the most clinically important observations was that a substantial proportion of patients who ultimately required cardiac intervention were asymptomatic at the time of return to care. Examples included patients with severe subaortic stenosis with peak gradients exceeding 80 mmHg, a restrictive ventricular septal defect with newly developed aortic regurgitation, a large atrial septal defect with right heart dilation, and moderate re-coarctation following prior repair – all identified only through imaging at the index return visit. In contrast, other patients presented with advanced disease, including acute heart failure with severe left ventricular dysfunction, cyanosis necessitating urgent transplant evaluation, infective endocarditis, and an aortic root aneurysm complicated by severe left ventricular dysfunction that ultimately resulted in death. These two distinct presentation patterns, silent progression and acute decompensation, reflect the same underlying problem: when longitudinal surveillance is interrupted, the natural course of CHD becomes unpredictable, and many of the resulting presentations are higher-acuity, higher-risk, and largely preventable.^11,25^

Notably, nearly two-thirds of patients who RTC had no documented reason for re-engagement. This finding has not been well characterized in the literature and warrants further study. Possible explanations include unrecorded family-driven re-engagement, primary care referrals, school or sports requirements, and reactivation of insurance coverage. Whatever the underlying drivers, this pattern suggests that re-engagement is often opportunistic rather than systematic, reinforcing the need for proactive outreach mechanisms rather than reliance on patient-initiated return.

These findings also have important implications for patient and family education and for transition of care. The multivariable association between older age and GIC is consistent with literature describing the adolescent and young adult period as a particularly high-risk window for disengagement.^10,12,25,28,34,36–40^ The absence of symptoms is frequently misinterpreted as the absence of disease, especially among adolescents and young adults who may have limited understanding of their CHD diagnosis, prior surgical history, and long-term prognosis.^11,12,38,41^ Reinforcing the concept that being asymptomatic is not equivalent to being disease-free, particularly during the transition from pediatric to adult-centered care, is essential to preventing the late, high-acuity presentations observed in this cohort.

An additional, often under-recognized contributor to GIC is variability in clinician-driven follow-up recommendations. Differences in perceived risk, especially for patients with simple CHD, may yield follow-up intervals that fail to account for the potential for late complications and silent progression. Standardized, evidence-informed follow-up pathways stratified by anatomic complexity, paired with clinician education to reduce inter-provider variability, represent a tractable system-level intervention.^4,5^

Addressing GIC will require a shift from reactive to proactive models of care. Increasing follow-up frequency alone is unlikely to address the underlying drivers of disengagement and may in fact lead to more disengagement and drive up costs for patients. What is needed is a comprehensive model that integrates longitudinal surveillance with systematic identification of patient needs. Within this framework, the routine incorporation of patient-reported outcome measures into clinical workflows provides real-time insights across multiple domains, including insurance stability, financial strain, mental health, caregiver support, transition readiness, and ancillary needs such as dental care.^31^ Equally important is ensuring that responses to patient-reported outcome measures are linked to actionable outputs, including clear referral pathways, care coordination resources, and targeted support services. Within a value-based care framework, this approach allows alignment of resources with patient-specific risk and may meaningfully reduce GIC and its downstream consequences. In parallel, emerging analytic approaches, including artificial intelligence and large language models, offer the potential to leverage both structured and unstructured EMR data to detect patterns associated with GIC, such as missed appointments, declining engagement, and evolving social needs, thereby enabling earlier, more anticipatory intervention.

## Limitations

This study should be interpreted in the context of several limitations. It was conducted at a single tertiary center, which may limit generalizability to other health systems with different patient populations, referral patterns, and care models. Although extensive chart review and external record evaluation were performed, some patients categorized as having GIC or LTFU may have received care outside of accessible systems, leading to potential misclassification. Conversely, patients categorized as having an appropriate care pathway may still have experienced clinically relevant disease progression that were not captured within our definition, particularly given variability in clinician follow-up recommendations and transitions across care systems. Identification of GIC contributors relied on available documentation, which is inherently incomplete and variable, and likely underestimates the true burden of insurance and social challenges. In addition, the retrospective design limits the ability to establish causality between GIC and adverse outcomes, and unmeasured confounding factors may influence both care continuity and clinical trajectories. Despite these limitations, the use of detailed chart review and the linkage of GIC to clinical outcomes provides a more granular understanding of care discontinuity than prior studies that rely solely on structured data.

## Conclusions

GIC remain prevalent in patients with surgically repaired CHD and carry meaningful clinical consequences, including the need for semi-urgent or urgent intervention and associated morbidity and mortality. These gaps occur across the full spectrum of anatomic complexity, including patients with simple CHD, and are most often driven by insurance instability and social vulnerability rather than clinical factors. Many of the adverse outcomes observed may be preventable with consistent longitudinal care. Addressing GIC will require a shift from reactive to proactive care models that integrate standardized follow-up pathways, systematic assessment of patient-level needs, and emerging analytical tools to identify at-risk patients before disengagement occurs.

## Data Availability

The data that support the findings of this study are not publicly available due to patient privacy and institutional restrictions but are available from the corresponding author upon reasonable request and with appropriate institutional approvals.

## Acknowledgments

The authors acknowledge the assistance of Editorial Services of Nemours Children’s Health.

## REFERENCES

1. Martin SS, Aday AW, Allen NB, Almarzooq ZI, Anderson CAM, Arora P, Avery CL, Baker-Smith CM, Bansal N, Beaton AZ, et al. Correction to: 2025 Heart disease and stroke statistics: a report of US and global data from the American Heart Association. Circulation. 2025;151:e1096. doi:10.1161/CIR.0000000000001345

2. Martin SS, Aday AW, Almarzooq ZI, Anderson CAM, Arora P, Avery CL, Baker-Smith CM, Barone Gibbs B, Beaton AZ, Boehme AK, et al. Correction to: 2024 Heart disease and stroke statistics: a report of US and global data from the American Heart Association. Circulation. 2025;151:e1095. doi:10.1161/CIR.0000000000001344

3. Mandalenakis Z, Giang KW, Eriksson P, Liden H, Synnergren M, Wåhlander H, Fedchenko M, Rosengren A, Dellborg M. Survival in children with congenital heart disease: have we reached a peak at 97%?. J Am Heart Assoc. 2020;9:e017704. doi:10.1161/JAHA.120.017704

4. Gurvitz M, Krieger EV, Fuller S, Davis LL, Kittleson MM, Aboulhosn JA, Bradley EA, Buber J, Daniels CJ, Dimopoulos K, et al. Correction to: 2025 ACC/AHA/HRS/ISACHD/SCAI Guideline for the management of adults with congenital heart disease: a report of the American College of Cardiology/American Heart Association Joint Committee on Clinical Practice Guidelines. Circulation. 2026;153:e1115. doi:10.1161/CIR.0000000000001432

5. Stout KK, Daniels CJ, Aboulhosn JA, Bozkurt B, Broberg CS, Colman JM, Crumb SR, Dearani JA, Fuller S, Gurvitz M, et al. 2018 AHA/ACC Guideline for the management of adults with congenital heart disease: executive summary: a report of the American College of Cardiology/American Heart Association Task Force on Clinical Practice Guidelines. Circulation. 2019;139:e637–e697. doi:10.1161/CIR.0000000000000602

6. Baumgartner H, De Backer J. The ESC clinical practice guidelines for the management of adult congenital heart disease 2020. Eur Heart J. 2020;41:4153–4154. doi:10.1093/eurheartj/ehaa701

7. Marelli A, Beauchesne L, Colman J, Ducas R, Grewal J, Keir M, Khairy P, Oechslin E, Therrien J, Vonder Muhll IF, et al. Canadian Cardiovascular Society 2022 guidelines for cardiovascular interventions in adults with congenital heart disease. Can J Cardiol. 2022;38:862–896. doi:10.1016/j.cjca.2022.03.021

8. Zaidi AH, Saleeb SF, Gurvitz M, Bucholz E, Gauvreau K, Jenkins KJ, de Ferranti SD. Social determinants of health including Child Opportunity Index leading to gaps in care for patients with significant congenital heart disease. J Am Heart Assoc. 2024;13:e028883. doi:10.1161/JAHA.122.028883

9. Bucholz EM, Sleeper LA, Newburger JW. Neighborhood socioeconomic status and outcomes following the Norwood procedure: an analysis of the Pediatric Heart Network single ventricle reconstruction trial public data set. J Am Heart Assoc. 2018;7:e007065. doi:10.1161/JAHA.117.007065

10. Yeung E, Kay J, Roosevelt GE, Brandon M, Yetman AT. Lapse of care as a predictor for morbidity in adults with congenital heart disease. Int J Cardiol. 2008;125:62–65. doi:10.1016/j.ijcard.2007.02.023

11. Gurvitz M, Valente AM, Broberg C, Cook S, Stout K, Kay J, Ting J, Kuehl K, Earing M, Webb G, et al. Prevalence and predictors of gaps in care among adult congenital heart disease patients: HEART-ACHD (The Health, Education, and Access Research Trial). J Am Coll Cardiol. 2013;61:2180–2184. doi:10.1016/j.jacc.2013.02.048

12. Gurvitz MZ, Inkelas M, Lee M, Stout K, Escarce J, Chang RK. Changes in hospitalization patterns among patients with congenital heart disease during the transition from adolescence to adulthood. J Am Coll Cardiol. 2007;49:875–882. doi:10.1016/j.jacc.2006.09.051

13. Zaidi AH, Alberts A, Chowdhury D, Beaty C, Brewer B, Chen MH, de Ferranti SD. Trends in gaps of care for patients with congenital heart disease: implications for social determinants of health and Child Opportunity Index. J Am Heart Assoc. 2024;13:e034796. doi:10.1161/JAHA.124.034796

14. Mylotte D, Pilote L, Ionescu-Ittu R, Abrahamowicz M, Khairy P, Therrien J, Mackie AS, Marelli A. Specialized adult congenital heart disease care: the impact of policy on mortality. Circulation. 2014;129:1804–1812. doi:10.1161/CIRCULATIONAHA.113.005817

15. Mackie AS, Ionescu-Ittu R, Therrien J, Pilote L, Abrahamowicz M, Marelli AJ. Children and adults with congenital heart disease lost to follow-up: who and when?. Circulation. 2009;120:302–309. doi:10.1161/CIRCULATIONAHA.108.839464

16. Mackie AS, Rempel GR, Rankin KN, Nicholas D, Magill-Evans J. Risk factors for loss to follow-up among children and young adults with congenital heart disease. Cardiol Young. 2012;22:307–315. doi:10.1017/S104795111100148X

17. Miao Q, Dunn S, Wen SW, Lougheed J, Reszel J, Lavin Venegas C, Walker M. Neighbourhood maternal socioeconomic status indicators and risk of congenital heart disease. BMC Pregnancy Childbirth. 2021;21:72. doi:10.1186/s12884-020-03512-8

18. Taylor LC, Burke B, Donohue JE, Yu S, Hirsch-Romano JC, Ohye RG, Goldberg CS. Risk factors for interstage mortality following the Norwood procedure: impact of sociodemographic factors. Pediatr Cardiol. 2016;37:68–75. doi:10.1007/s00246-015-1241-2

19. Burch PT, Gerstenberger E, Ravishankar C, Hehir DA, Davies RR, Colan SD, Sleeper LA, Newburger JW, Clabby ML, Williams IA, et al. Longitudinal assessment of growth in hypoplastic left heart syndrome: results from the single ventricle reconstruction trial. J Am Heart Assoc. 2014;3:e000079. doi:10.1161/JAHA.114.000079

20. Ravishankar C, Gerstenberger E, Sleeper LA, Atz AM, Affolter JT, Bradley TJ, Gaynor JW, Goldstein BH, Henderson HT, Jacobs JP, et al. Factors affecting Fontan length of stay: results from the single ventricle reconstruction trial. J Thorac Cardiovasc Surg. 2016;151:669–675.e1. doi:10.1016/j.jtcvs.2015.09.061

21. Xiang L, Su Z, Liu Y, Huang Y, Zhang X, Li S, Zhang H. Impact of family socioeconomic status on health-related quality of life in children with critical congenital heart disease. J Am Heart Assoc. 2019;8:e010616. doi:10.1161/JAHA.118.010616

22. Connor JA, Kline NE, Mott S, Harris SK, Jenkins KJ. The meaning of cost for families of children with congenital heart disease. J Pediatr Health Care. 2010;24:318–25. doi:10.1016/j.pedhc.2009.09.002

23. Connor JA, Gauvreau K, Jenkins KJ. Factors associated with increased resource utilization for congenital heart disease. Pediatrics. 2005;116:689–695. doi:10.1542/peds.2004-2071

24. Sistino JJ, Ellis C. Effects of health disparities on survival after neonatal heart surgery: why should racial, ethnic, gender, and socioeconomic status be included in the risk analysis?. J Extra Corpor Technol. 2011;43:232–235.

25. Moore JA, Sheth SS, Lam WW, Alexander AJ, Shabosky JC, Espaillat A, Lovick DK, Broussard NS, Dyer KJ, Lopez KN. Hope is no plan: uncovering actively missing transition-aged youth with congenital heart disease. Pediatr Cardiol. 2022;43:1046–1053. doi:10.1007/s00246-022-02823-1

26. Davey B, Sinha R, Lee JH, Gauthier M, Flores G. Social determinants of health and outcomes for children and adults with congenital heart disease: a systematic review. Pediatr Res. 2021;89:275–294. doi:10.1038/s41390-020-01196-6

27. Tillman AR, Colborn KL, Scott KA, Davidson AJ, Khanna A, Kao D, McKenzie L, Ong T, Rausch CM, Duca LM, et al. Associations between socioeconomic context and congenital heart disease related outcomes in adolescents and adults. Am J Cardiol. 2021;139:105–115. doi:10.1016/j.amjcard.2020.10.040

28. Ko JM, Yanek LR, Cedars AM. Factors associated with a lower chance of having gaps in care in adult congenital heart disease. Cardiol Young. 2021;31:1576–1581. doi:10.1017/S1047951121000524

29. Moons P, Hilderson D, Van Deyk K. Implementation of transition programs can prevent another lost generation of patients with congenital heart disease. Eur J Cardiovasc Nurs. 2008;7:259–263. doi:10.1016/j.ejcnurse.2008.10.001

30. Moons P, De Volder E, Budts W, De Geest S, Elen J, Waeytens K, Gewillig M. What do adult patients with congenital heart disease know about their disease, treatment, and prevention of complications? A call for structured patient education. Heart. 2001;86:74–80. doi:10.1136/heart.86.1.74

31. Chowdhury D, Johnson JN, Baker-Smith CM, Jaquiss RDB, Mahendran AK, Curren V, Bhat A, Patel A, Marshall AC, Fuller S, et al. Health care policy and congenital heart disease: 2020 focus on our 2030 future. J Am Heart Assoc. 2021;10:e020605. doi:10.1161/JAHA.120.020605

32. Videbæk J, Laursen HB, Olsen M, Høfsten DE, Johnsen SP. Long-term nationwide follow-up study of simple congenital heart disease diagnosed in otherwise healthy children. Circulation. 2016;133:474–483. doi:10.1161/CIRCULATIONAHA.115.017226

33. Noelke C, McArdle N, DeVoe B, Leonardos M, Lu Y, Ressler RW, Acevedo-Garcia D. COI 3.0 Technical Documentation How we built the Child Opportunity Index 3.0. diversitydatakids.org. https://www.diversitydatakids.org/research-library/research-report/coi-30-technical-documentation. Accessed July 26, 2024.

34. Helm PC, Kempert S, Körten MA, Lesch W, Specht K, Bauer UMM. Congenital heart disease patients’ and parents’ perception of disease-specific knowledge: Health and impairments in everyday life. Congenit Heart Dis. 2018;13:377–383. doi:10.1111/chd.12581

35. Hoffman JI, Kaplan S, Liberthson RR. Prevalence of congenital heart disease. Am Heart J. 2004;147:425–439. doi:10.1016/j.ahj.2003.05.003

36. Scott J, Agarwala A, Baker-Smith CM, Feinstein MJ, Jakubowski K, Kaar J, Parekh N, Patel KV, Stephens J; American Heart Association Prevention Science Committee of the Council on Epidemiology and Prevention and Council on Cardiovascular and Stroke Nursing, et al. Cardiovascular health in the transition from adolescence to emerging adulthood: a scientific statement from the American Heart Association. J Am Heart Assoc. 2025;14:e039239. doi:10.1161/JAHA.124.039239

37. Salciccioli KB, Salemi JL, Broda CR, Lopez KN. Disparities in insurance coverage among hospitalized adult congenital heart disease patients before and after the Affordable Care Act. Birth Defects Res. 2021;113:644–659. doi:10.1002/bdr2.1878

38. Chong LSH, Fitzgerald DA, Craig JC, Manera KE, Hanson CS, Celermajer D, Ayer J, Kasparian NA, Tong A. Children’s experiences of congenital heart disease: a systematic review of qualitative studies. Eur J Pediatr. 2018;177:319–336. doi:10.1007/s00431-017-3081-y

39. Knauth Meadows A, Bosco V, Tong E, Fernandes S, Saidi A. Transition and transfer from pediatric to adult care of young adults with complex congenital heart disease. Curr Cardiol Rep. 2009;11:291–297. doi:10.1007/s11886-009-0042-8

40. Reiss JG, Gibson RW, Walker LR. Health care transition: youth, family, and provider perspectives. Pediatrics. 2005;115:112–120. doi:10.1542/peds.2004-1321

41. Lesch W, Specht K, Lux A, Frey M, Utens E, Bauer U. Disease-specific knowledge and information preferences of young patients with congenital heart disease. Cardiol Young. 2014;24:321–330. doi:10.1017/S1047951113000413

